# Impaired Neutralization of SARS-CoV-2 Including Omicron Variants after COVID-19 mRNA Booster Immunization under Methotrexate Therapy

**DOI:** 10.1101/2022.06.11.22276272

**Authors:** Elisa Habermann, Lutz Gieselmann, Pinkus Tober-Lau, Alexander ten Hagen, Fredrik N Albach, Jan Zernicke, Elvin Ahmadov, Amanthi Nadira Arumahandi de Silva, Leonie Maria Frommert, Jens Klotsche, Florian Kurth, Leif Erik Sander, Gerd Burmester, Florian Klein, Robert Biesen

## Abstract

**Objective:** To determine the immediate need for a fourth COVID-19 vaccination based on the neutralizing capacity in patients on methotrexate (MTX) therapy after mRNA booster immunization.

**Methods:** In this observational cohort study, neutralizing serum activity against SARS-CoV-2 wildtype (Wu01) and variant of concern (VOC) Omicron BA.1 and BA.2 were assessed by pseudovirus neutralization assay before, 4 and 12 weeks after mRNA booster immunization in 50 rheumatic patients on MTX, 26 of whom paused the medication. 44 non-immunosuppressed persons (NIP) served as control group.

**Results:** While the neutralizing serum activity against SARS-CoV-2 Wu01 and Omicron variants increased 67-to 73-fold in the NIP after booster vaccination, the serum activity in patients receiving MTX increased only 20-to 23-fold. As a result, significantly lower neutralizing capacities were measured in patients on MTX compared to the NIP at week 4. Patients who continued MTX treatment during vaccination had significantly lower neutralizing serum titres against all three virus strains at week 4 and 12 compared to patients who paused MTX and the control group, except for BA.2 at week 12. Patients who paused MTX reached comparably high neutralization titres as the NIP, except for Wu01 at week 12. Neutralization of omicron variants was significantly lower in comparison to wildtype in both groups.

**Conclusion:** Patients pausing MTX showed a similar vaccine response to NIP. Patients who continued MTX demonstrated an impaired booster response indicating a potential benefit of a second booster vaccination.

## INTRODUCTION

SARS-CoV-2 has caused at least 520 million confirmed infections and 6.25 million deaths worldwide by June 2022 [1]. Over time, naturally occurring mutations alter the genome of SARS-CoV-2. If the evolved virus variants show increased transmissibility and/or virulence, disease severity and escape from humoral immunity, they are designated as variants of concern (VOC). One of these VOCs, which is globally prevalent in early 2022, is the Omicron variant and its sublineages BA.1 and BA.2 [1]. It displays an unusually high number of mutations in the receptor-binding (RBD) or N-terminal domain of the viral spike (S) protein. Some of these mutations were already identified in other VOCs and are associated with increased susceptibility and escape from neutralizing antibody responses [2]. Fortunately, the T cell reactivity against the Omicron variant is not reduced after basic immunization [3] and booster vaccination with wild-type spike mRNA induces robust levels of neutralizing serum activity against the Omicron variant [4]. Thus, these vaccines continue to provide protection against severe disease [5,6].

Various immunosuppressants reduce the immune response after COVID-19 vaccination [7,8]. Methotrexate (MTX) is the most commonly prescribed disease-modifying antirheumatic drug in the world [9]. MTX reduces the humoral vaccination response and CD8+ T-cell activation after second vaccination against COVID-19 [10]. Pausing MTX therapy 10 or 14 days after both vaccinations of the basic immunization against COVID-19 significantly improves the production of neutralizing antibodies [11,12]. To our knowledge, the effect of MTX on booster vaccination has only been reported once. In this study, MTX patients showed no reduction in vaccine antibodies against the SARS-CoV-2 Wu01, as all 269 patients paused therapy for 2 weeks after booster with CoronaVac vaccine (Sinovac Biotech) [13]. The effect of continued MTX on neutralization activity, especially regarding the Omicron variant, remains to be elucidated.

International and national authorities and commissions worldwide have recommended a fourth COVID-19 vaccination for immunocompromised patients [14–17]. As there is no data to document whether this recommendation is also justified for MTX patients, the aim of this work was to compare the neutralizing capacity against SARS-CoV2 and variants in MTX patients after COVID-19 booster vaccination with that of non-immunosuppressed persons.

## METHODS

### Study design and participants

This is the continuation of our recently published sub-analysis of the VACCIMMUN study, which investigated the factors influencing the humoral immune response of a COVID-19 basic immunization in MTX patients [11]. Blood samples were collected under identical inclusion and exclusion criteria from MTX patients and NIP shortly before, 4 and 12 weeks after an mRNA booster vaccination in the period from July 2021 to March 2022. Samples from individuals who had a COVID-19 infection prior to one of the blood collections were excluded. The patients provided information regarding medical history including COVID-19 vaccination status and/or infection and immunosuppressive therapy directly.

MTX patients were asked at week 4 post booster vaccination whether MTX was paused and for how long. Instructions to continue or withhold MTX were not given in this study but observed as part of it.

### Laboratory analyses

#### SARS-CoV-2 pseudovirus constructs

The nucleotide sequence of expression plasmids encoding all SARS-CoV-2 spike proteins was codon-optimized. The SARS-CoV-2 pseudovirus expressing the Wu01 spike protein (EPI_ISL_40671) was generated using expression plasmids that include a C-terminal deletion of 21 cytoplasmatic amino acids to achieve enhanced pseudovirus titers. Expression plasmids encoding the spike proteins of Omicron sublineage (BA.1 and BA.2) were generated by assembly and cloning of codon-optimized overlapping gene fragments (Thermo Fisher) into the pCDNA3.1/V5-HisTOPO vector (Thermo Fisher) using the NEBuilder Hifi DNA Assembly Kit (New England Biolabs). Expression plasmids for the Omicron sublineage included the following amino acid changes relative to Wu01:

Lineage B.1.1.529, sublineage BA.1: A67V, Δ69-70, T95I, G142D, Δ143-145, N211I, Δ212, ins215EPE, G339D, S371L, S373P, S375F, K417N, N440K, G446S, S477N, T478K, E484A, Q493R, G496S, Q498R, N501Y, Y505H, T547K, D614G, H655Y, N679K, P681H, N764K, D796Y, N856K, Q954H, N969K, and L981F.

Lineage B.1.1.529, sublineage BA.2: T19I, Δ24-26, A27S, A67V, G142D, V213G, G339D, S371F, S373P, S375F, T376A, D405N, R408S, K417N, N440K, S477N, T478K, E484A, Q493R, Q498R, N501Y, Y505H, D614G, H655Y, N679K, P681H, N764K, D796Y, Q954H, N969K.

Sequences of all expression plasmids were verified by Sanger Sequencing.

#### SARS-CoV-2 pseudovirus neutralization assays

SARS-CoV-2 pseudoviruses were generated by co-transfection of individual plasmids encoding HIV Tat, HIV Gag/Pol, HIV Rev, luciferase followed by an IRES and ZsGreen, and the SARS-CoV-2 spike protein (Wu01, BA.1 and BA.2) in adherent HEK 293T cells using the FuGENE 6 Transfection Reagent (Promega). Cell culture supernatants containing pseudovirus particles were harvested 48-72h after transfection, centrifuged, filtered using a 0.45 µm filter, and stored at -80°C till use. Titration of the pseudoviruses was performed by infection of HEK293T cells expressing human ACE2 (Crawford et al., 2020) at 37°C and 5% CO2. After an incubation period of 48h, luciferase activity was determined by addition of luciferin/lysis buffer (10 mM MgCl2, 0.3 mM ATP, 0.5 mM Coenzyme A, 17 mM IGEPAL (all Sigma-Aldrich), and 1 mM D-Luciferin (GoldBio) in Tris-HCL) using a microplate reader (Berthold). For neutralization assays, a virus dilution with a relative luminescence unit (RLU) of approximately 1,000-fold in infected cells versus non-infected cells was selected.

Before usage, serum samples of study participants were inactivated at 56°C for 40 min. For determination of the serum neutralizing activity, three-fold serial dilutions of samples (starting dilution at 1:10) in cell culture medium were co-incubated with pseudovirus supernatants for 1h at 37°C and 293T-ACE2 cells were added. After incubation for 48h at 37 °C and 5 % CO2, luciferase activity was determined using the luciferin/lysis buffer. The background RLUs of non-infected cells was subtracted and the 50% inhibitory serum dilution (ID50) that resulted in a 50% reduction of signal compared to the virus-infected untreated control was determined using a non-linear fit model to plot an agonist vs. normalized dose response curve with variable slope using the least squares fitting method in GraphPad Prism 7.0 (GraphPad). All serum samples were tested in duplicates.

#### Statistical analysis

Descriptive statistics included mean with SD, geometric mean with 95% CI and absolute and relative frequencies. The unpaired t-test with Welch’s correction was performed to compare continuously distributed variables and the binomial test for parts of a whole for binary data in Table 1.

**Table 1:** Characteristics of patients and controls

| Characteristics | MTX n=50 | NIP n=44 | P value |
| --- | --- | --- | --- |
| age, mean (SD) | 61.68 (12.4) | 61.9 (21.5) | 0.946 |
| female, n (%) | 40 (80) | 29 (65.9) | 0.339 |
| BMI mean, (SD) | 25.7 (4.0) | 25.77 (4.5) | 0.927 |
| Booster vaccination |  |  |  |
| BNT162b2, n (%) | 38 (76) | 44 (100) | *0.032 |
| mRNA-1273, n (%) | 12 (24) | 0 (0) |  |
| Time between blood sampling and vaccination |  |  |  |
| days between first visit and booster (pre booster), mean (range) | 10 (0-30) | 7 (0-36) | 0.089 |
| days between booster and second visit (week 4), mean (range) | 31 (25-53) | 27 (17-42) | *0,001 |
| days between booster and third visit (week 12), mean (range) | 88 (76-129) | 97 (78-124) | *0,009 |
| Rheumatic diagnosis |  |  |  |
| Rheumatoid arthritis, n (%) | 36 (72) | / |  |
| Psoriatic arthritis, n (%) | 7 (14) | / |  |
| other, n (%)* | 7 (14) | / |  |
| Medication |  |  |  |
| MTX-mono, n (%) | 15 (30) | / |  |
| MTX+prednisolone, n (%) | 12 (24) | / |  |
| MTX+TNF-α-inhibitor, n (%) | 11 (22) | / | |
| MTX+TNF-α-inhibitor+prednisolone, n (%) | 6 (12) | / | |
| MTX+other, n (%)** | 6 (12) | / |  |
| MTX-dose in mg/week, mean (SD) | 12.5 (4.2) | / |  |
| Prednisolone in mg/d, mean (SD) | 3.8 (3.3) | / |  |
| MTX regimen |  |  |  |
| MTX continued, n (%) | 24 (48) | / |  |
| MTX pause, n (%) | 26 (52) | / |  |
| duration of hold (days), mean (SD) | 19.9 (7.7) | / |  |
| duration of hold before vaccine (days), mean (SD) | 7.6 (3.9) | / |  |
| duration of hold after vaccine (days), mean (SD) | 12.4 (7.1) | / |  |

\*Takayasu arteritis, 2x Axial spondylarthritis (Ankylosing spondylitis), Dermatomyositis/polymyositis, Polymyalgia rheumatica, Blistering dermatitis, Systemic sclerosis
\*\*2 x Leflunomide; IL-12/IL-23-Inhibitor; Prednisolone/Immunglobulins; Hydroxychloroquine; IL-17-Inhibitor

Neutralizing antibody levels were not normally distributed and therefore differences between defined groups (e.g., MTX vs NIP or MTX pause vs non-pause) were analyzed using the Mann-Whitney U test (MWUT). GraphPad Prism 9.3.0 was used for all statistical analyses.

#### Patient and public involvement

There was no patient or public involvement in the designing of this study.

## RESULTS

### Patient characteristics

Of 65 people on MTX therapy with blood samples taken four weeks after booster vaccination 15 had to be excluded due to unacceptable immunosuppressive comedication known to significantly decrease vaccination response, such as rituximab [7]. One participant had to be excluded from the study 12 weeks after vaccination due to a COVID-19 infection.

Neutralizing serum activity of patients on MTX against SARS-CoV-2 variants Wuhan, Omicron BA.1, and BA.2 was compared with NIP before, 4, and 12 weeks after COVID-19 mRNA booster immunization (figure 1).

**Figure 1:**
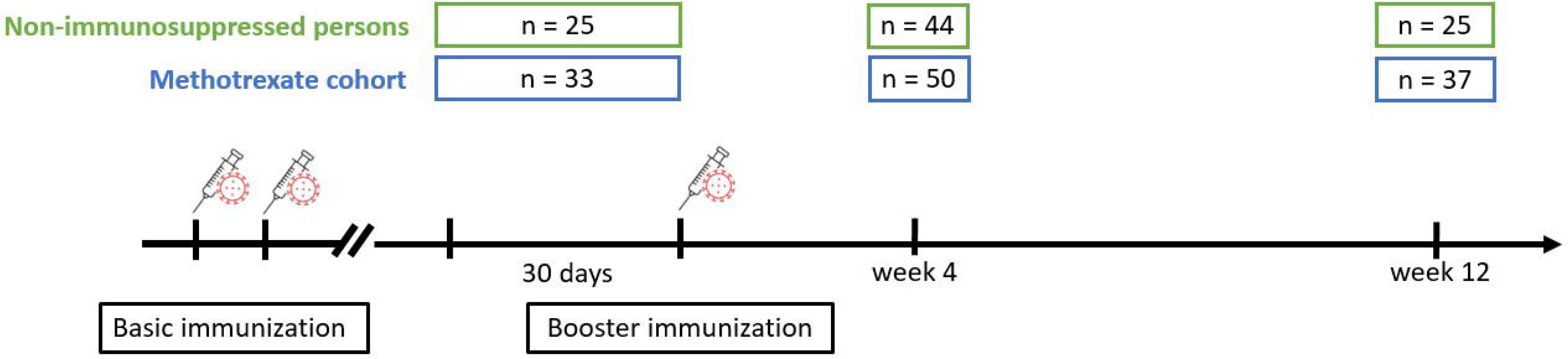
Observational study design around booster vaccination including number of participants at each visit.

There were no significant differences between the groups regarding age, gender and body mass index (table 1). However, both groups were significantly different regarding the vaccines administered, since 100% of NIP received the BNT162b2 vaccine while 24% of MTX patients were vaccinated with the mRNA-1273 vaccine. Detailed clinical characterization of the MTX and NIP control cohort can be found in table 1.

### Impaired SARS-CoV-2 neutralizing activity in sera from MTX patients

In the following, we compare serum neutralizing capacity of MTX patients with NIP against different virus variants over the three visits. We discovered that the neutralizing serum activity against the two SARS-CoV-2 variants Wu01 and Omicron (sublineages BA.1 and BA.2) was reduced in MTX patients compared to the control group (Figure 2). Furthermore, the neutralizing activity against the Omicron sublineages was significantly lower than against Wu01.

**Figure 2:**
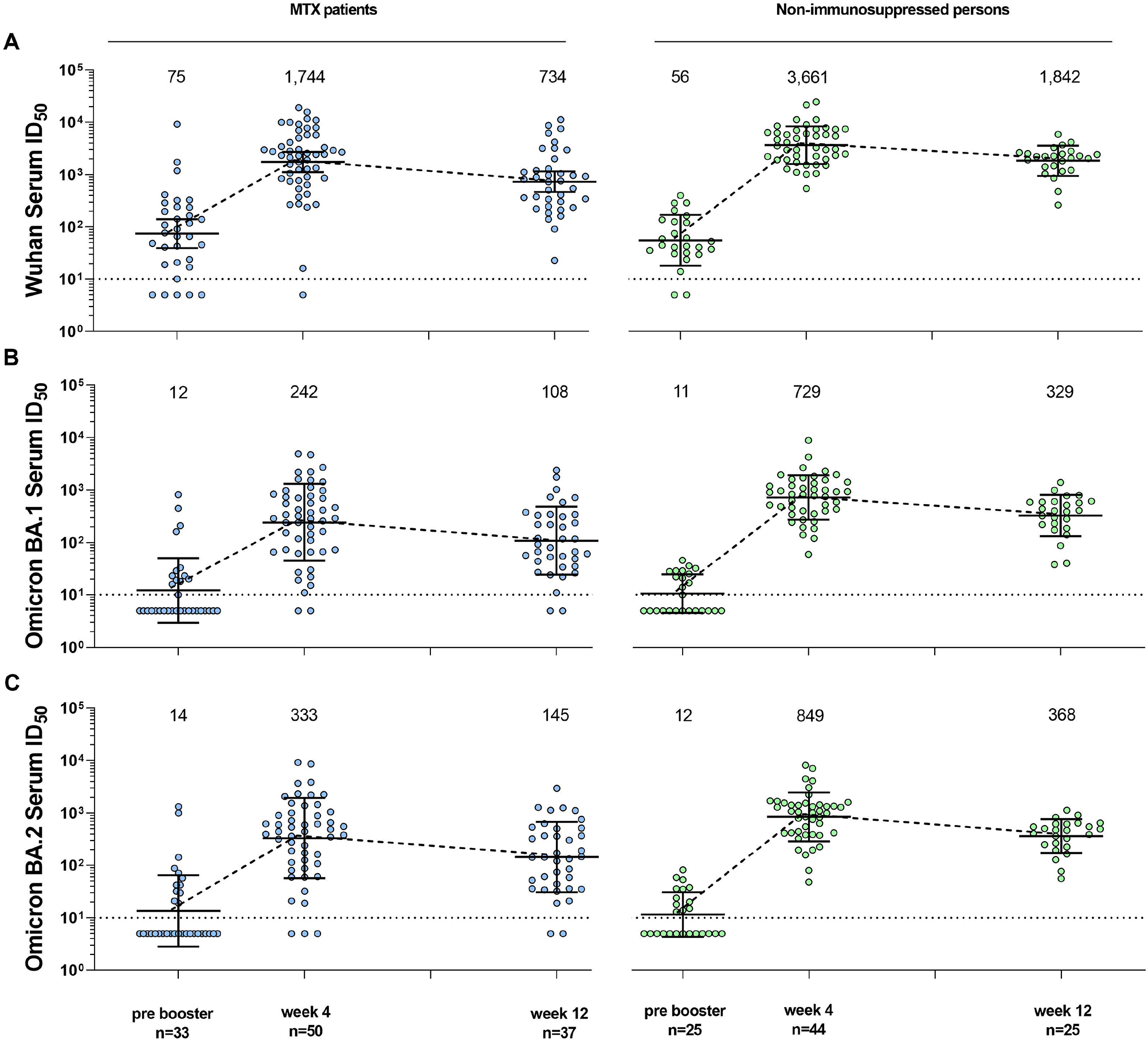
Neutralizing serum activity against SARS-CoV-2 Wu01 and Omicron variant (sublineages BA.1 and BA.2) in patients with methotrexate (left side) versus health care workers (right side) before, 4 weeks and 12 weeks after mRNA-Booster vaccination. 50 % inhibitory serum dilutions (ID_50_s) were determined by pseudovirus neutralization assays. Dot plots and numbers above the graph illustrate the geometric mean ID_50_ and error bars indicate the 95 % confidence intervals. Dotted black lines display the lower limit of quantification (LLOQ) of the neutralization assay (ID_50_ of 10). ID_50_s below the LLOQ (ID_50_ = 10) were assigned to half the LLOQ (ID_50_ = 5).

#### Impaired neutralization activity against Wuhan variant in MTX patients

Serum neutralizing activity against the Wu01 variant before booster vaccination was not significantly different between MTX patients and controls (p=0.424, MWUT, Figure 2A).

While Wu01-neutralizing serum titers in NIP increased 67-fold (to a geometric mean ID_50_ of 3,735) 4 weeks after vaccination, in MTX patients titers increased only 23 fold (to a geometric mean ID_50_ of 1,744). This resulted in a significantly lower neutralizing serum activity against the Wu01 variant in the MTX patients 4 weeks after vaccination than in the controls (p=0.015, MWUT).

Serum ID_50_s of both groups decreased to about half from week 4 to week 12 (to a geometric mean ID_50_ of 734 for MTX patients and 1842 for NIP, respectively). At week 12, the Serum ID_50_s were also significantly lower for the MTX patients than for the NIPs (p=0.001, MWUT).

#### Impaired neutralization activity against the Omicron BA.1 in MTX patients

Serum neutralizing activity against Omicron BA.1 was not significantly different between MTX patients and controls before booster vaccination (p=0.657, MWUT, Figure 2B).

While the geometric mean ID_50_s in NIP increased 68-fold 4 weeks after booster immunization, the geometric mean ID_50_s in MTX patients increased only 20-fold. This resulted in a significantly lower serum neutralizing activity against Omicron BA.1 in MTX patients 4 weeks after booster immunization than in the controls (p<0.001, MWUT).

The geometric mean ID_50_s of both groups decreased to about half from week 4 to week 12. At week 12, serum neutralization activity was also significantly lower in the MTX patients than in the NIP (p=0.001, MWUT).

#### Impaired neutralization activity against the Omicron BA.2 in MTX patients

Serum neutralizing activity against Omicron BA.2 was not significantly different between MTX patients and controls before booster vaccination (p=0.838, MWUT, Figure 2C).

While the geometric mean ID_50_s in NIP increased 73-fold 4 weeks after booster immunization, the geometric mean ID_50_s in MTX patients increased only 23-fold. This resulted in a significantly lower serum neutralizing activity against Omicron BA.2 in the MTX patients 4 weeks after booster immunization than in the controls (p<0.001, MWUT).

The geometric mean ID_50_s of both groups decreased to about half from week 4 to week 12. Also at week 12, BA.2 neutralizing serum activity was significantly lower in the MTX patients than in the NIP (p=0.001, MWUT).

### Comparison of neutralizing activity of Wu01 against Omicron sublineages

The neutralizing capacities against the BA.1 sublineage were on average lower by a factor of 6.75 (6.25, 7.20, 6.79) in the MTX cohort and by a factor of 5.23 (5.09, 5.02, 5.59) in the NIP compared to Wu01 across the three time points.

The neutralizing capacities against the BA.2 sublineage were on average lower by a factor of 5.21 (5.36, 5.23, 5.06) in the MTX cohort and by a factor of 4.65 (4.66, 4.31, 5.00) in the NIP compared to Wu01 across the three time points.

Neutralizing activity against Wu01 was significantly higher in both study groups compared to both Omicron sublineages at week 4 and week 12 (always p<0.001). However, there was no significant difference between the Omicron sublineages BA.1 and BA.2 in both cohorts at any point in time (always p>0,05).

### Impact of MTX discontinuation on neutralizing capacities against SARS-CoV-2 variants

We and others have previously demonstrated that the humoral vaccination response after basic immunization against COVID-19 can be improved by pausing MTX [11,12]. Accordingly, we aimed to investigate whether this effect can also be observed 4 and / or 12 weeks after the booster immunization and whether the patients who paused MTX achieved similarly high neutralizing serum activity as untreated controls.

Of the 50 MTX patients whose neutralizing activity against SARS-CoV-2 variants was examined 4 weeks after mRNA booster immunization, 26 paused MTX and 24 did not.

Serum neutralization activity against the Wu01 variant at week 4 after booster vaccination (Figure 3A) was significantly lower in patients continuously taking MTX than in patients pausing their medication (p=0.008, MWUT) or in NIP (p<0.001, MWUT). Patients on MTX pause achieved comparably high neutralizing serum activity as controls. Interestingly, serum activity at week 12 was significantly lower in patients on MTX pause (p<0.001) and patients on continuous MTX (p=0.046) than in non-immunosuppressed controls.

**Figure 3:**
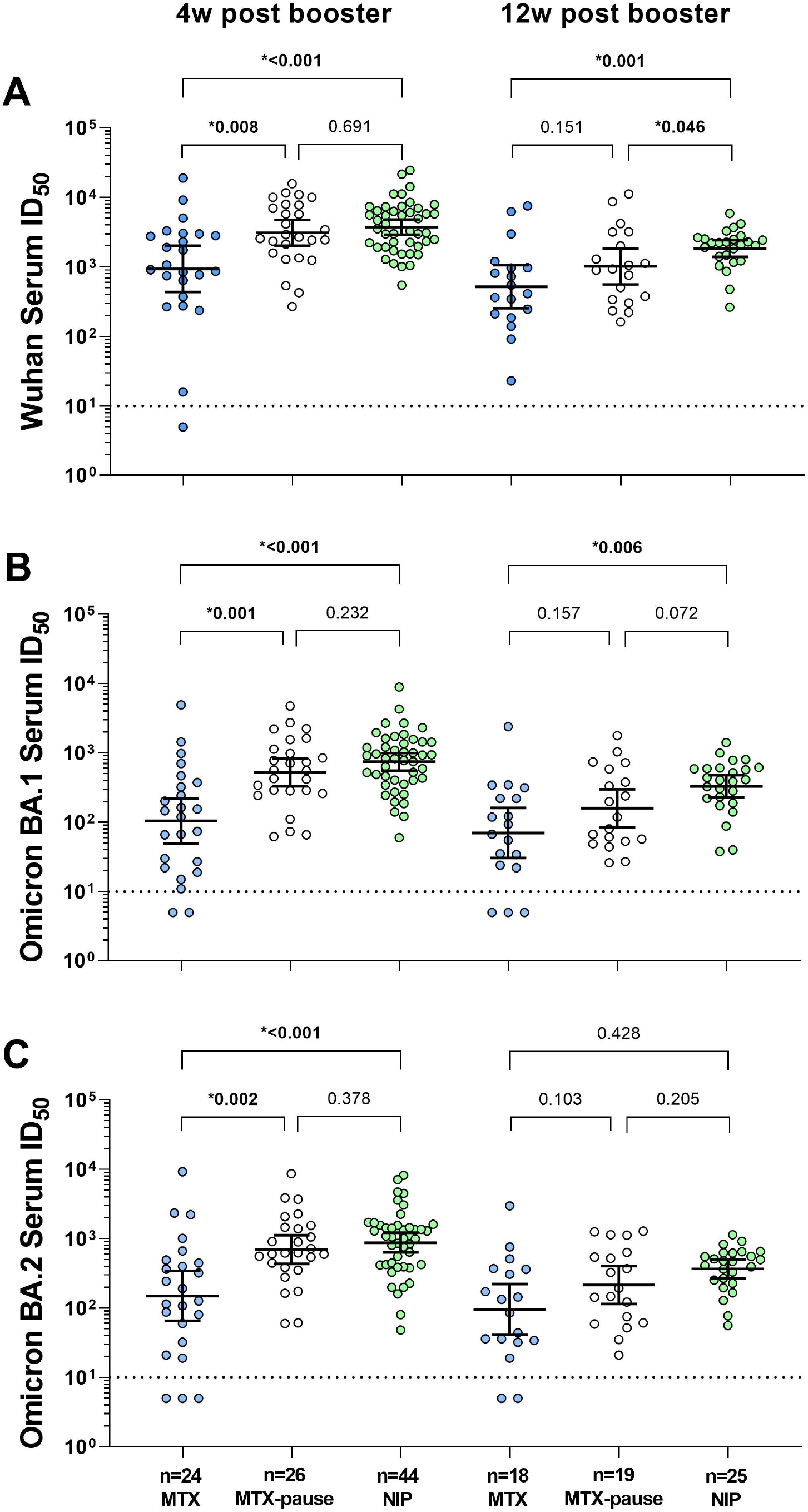
Serum neutralizing activity against Wu01, Omicron BA.1 and Omicron BA.2 4 and 12 weeks after mRNA booster vaccination in patients with and without MTX pause and healthcare workers. The p-values shown are based on the Mann-Whitney-U test, which was used to analyze the differences between the groups. 50 % inhibitory serum dilutions (ID_50_s) were determined by pseudovirus neutralization assays. Dot plots and numbers above the graph illustrate the geometric mean ID_50_ and error bars indicate the 95 % confidence intervals. Dotted black lines display the lower limit of quantification of the neutralization assay (ID_50_ of 10). ID_50_s below the LLOQ (ID_50_ = 10) were assigned to half the LLOQ (ID_50_ = 5).

Neutralization against Omicron BA.1 at week 4 after booster vaccination (Figure 3B) was significantly lower in patients taking continuous MTX in the MWUT (p<0.001) than in patients pausing MTX and in non-immunosuppressed controls (p<0.001). Again, patients with MTX pause achieved similar levels of serum neutralization activity compared to controls at week 4. At week 12, only neutralizing activity against BA.1 was significantly lower in patients taking continuous MTX than in NIP (p=0.006, MWUT), while there was no significant difference in patients pausing MTX (p=0.072, MWUT).

The neutralizing activity against Omicron BA.2 (Figure 3C) was also significantly lower at week 4 in patients taking continuous MTX in MWUT than in patients pausing MTX (p=0.002) and in non-immunosuppressed NIP (p<0.001). Again, at week 4, the neutralizing activity of paused MTX patients was not significantly different from that of controls. At week 12, the neutralizing activity against BA.2 was not significantly lower than in the non-immunosuppressed controls, neither in patients taking continuous MTX nor in patients with MTX pause.

### Other potential influencing factors on neutralizing capacity

Contrary to previous findings [11,12], there was no correlation between age and neutralizing capacity in the MTX cohort for any of the viral variants 4 and 12 weeks after booster vaccination (each p>0.05). The NIP showed a correlation between age and neutralizing capacity only for Wuhan at week 4 (p=0.037) and for Wuhan and Omicron BA.1 at week 12 (p=0.018 and p=0.017).

In addition, MTX dose had no effect on neutralization capacity in all MTX patients and in only the continuous MTX patients for any variant at any time point (each p>0.05).

## DISCUSSION

This is the first work demonstrating that MTX patients can also develop serum neutralization activity against the Omicron variant after a COVID-19 mRNA booster immunization. However, the extent depends largely on the pause of methotrexate. If patients continued to take MTX after vaccination, serum neutralization activity was significantly reduced. In contrast, patients who paused MTX after booster vaccination exhibited neutralizing capacities against all studied variants that were comparable to that of non-immunosuppressed individuals at week 4.

Before mRNA booster vaccination, MTX patients and controls had similar neutralizing activity. The overall increase and differences of neutralizing capacities against distinct virus variants after mRNA booster immunization measured in our cohort were comparable to previous studies [4,18,19]. The MTX patients exhibited a significantly lower increase than controls, resulting in markedly reduced neutralizing serum activity against all variants. We further show that discontinuation of MTX reversed the drug-mediated attenuation of the vaccination response. These results reconfirm that MTX attenuates the humoral vaccination response and is consistent with observations made after basic COVID-19 immunization [10,11] and influenza vaccination [20].

Although all patients on MTX pause had neutralizing antibodies against all variants and at all time points, it should be noted that the levels at week 12 after the booster were slightly lower than in NIP. This effect was only weakly significant in Wu01. The lack of significance in the Omicron sublineages may be due to a combination of a small number of cases and a flatter increase in neutralizing capacity against Omicron after booster vaccination.

This work has strengths and limitations. The strengths were a closely selected timeline for sample collection, a rigorous selection of MTX comedication [21], a follow up over 12 weeks, similar age and sex distribution among MTX patients and controls, and the reliable determination of serum neutralization activity. Various surrogate ELISA-based assays have been developed to evaluate SARS-CoV-2 neutralization capacity by measurement of binding inhibition of the receptor-binding domain (RBD) of the viral S protein to the cellular ACE2 receptor [22–24]. However, the informative value of these assays is limited by (i) cross-reactivity with neutralizing antibodies directed against other coronaviruses, (ii) the inability to measure synergistic neutralization activity of antibodies directed against distinct viral epitopes, (iii) the absence of non-RBD S protein epitopes, and (iiii) poor discrimination of high values. Therefore, virus neutralization assays are considered the gold standard for the determination of SARS-CoV-2 neutralizing serum activity [25]. Consequently, our study offers the highest available data validity.

Limitations include low number of cases, MTX pause recall bias in retrospective survey, and lack of systematic recording of disease activity and safety - although neither a disease flare nor a vaccine side effect was reported to us at week 4 or 12. We were unable to investigate T-cell function in this cohort, which leaves questions regarding this important aspect of immunogenicity unanswered.

In summary, defining the aim of booster vaccinations as the induction of neutralizing capacities against variants comparable to that of non-immunosuppressed persons, then there is a need for a fourth vaccination in patients that continued MTX during first booster vaccination. If possible, patients should pause MTX for further vaccinations.

## Data Availability

All data relevant to the study are included in the
article. Data are available on reasonable request.

## Key messages

### What is already known about this subject?

▸ In healthy individuals, mRNA-booster vaccination against COVID-19 induces a significantly improved neutralizing capacity against Omicron.
▸ Holding methotrexate (MTX) has shown to increase immunogenicity after COVID-19 vaccination.
▸ No previous studies have investigated the neutralizing capacity against Wuhan (Wu01) and Omicron after COVID-19 mRNA-booster vaccination in a MTX cohort.

### What does this study add?

▸ Most MTX patients developed detectable neutralizing capacities against Omicron sublineages after booster vaccination.
▸ Compared to individuals without immunosuppressive medication, MTX patients have a significantly lower neutralizing capacity against Wu01 and the Omicron sublineages after mRNA booster.
▸ Pausing MTX significantly improves the neutralizing capacity to a level comparable to individuals without immunosuppressive medication.

### How might this impact on clinical practice or future developments?

▸ Patients who continued MTX during the first booster vaccination should receive a second booster vaccination.
▸ Our data suggest that MTX patients should pause their medication for each booster vaccination.

## Acknowledgements

none

## Contributors

All authors contributed to the acquisition, analysis or interpretation of data and critical revision of the manuscript for important intellectual content. RB had full access to all the data in the study and takes responsibility for the integrity of the data and the accuracy of the data analysis. RB is responsible for the overall content as the guarantor. LES, FK and RB were involved in the study design. Sample collection was done by EH and AtH. Experiments and data analysis were performed by LG, EA, EH, and RB. EH, LG and RB were responsible for tables and figures. Data interpretation was done by all authors. Statistical analyses were done by EH, JK and RB. Writing of the manuscript were performed by EH, LG and RB. All authors were involved in critical proof reading of the manuscript.

## Funding

This work was supported by unconditional donations from Medac, Galapagos and Freunde und Förderer der Berliner Charité e.V.

This work was further supported by grants from COVIM: NaFoUniMedCovid19 (FKZ: 01KX2021) (to LES, FKl and FKu), the Federal Institute for Drugs and Medical Devices (V-2021.3 / 1503_68403 / 2021-2022) (to FKu and LES) and the German Center for Infection Research (DZIF) (to FKl), and the Deutsche Forschungsgemeinschaft (DFG) (CRC1310 to FKl and SFB-TR84 to LES).

### Competing interests

All authors declared no conflict of interests.

### Patient and public involvement

Patients and/or the public were not involved in the design, or conduct, or reporting, or dissemination plans of this research.

### Patient consent for publication

Not applicable.

### Ethics approval

This study was ethically approved by the Regional Office for Health and Social Affairs Berlin, Germany (21/0098-IV E 13). All patients provided written informed consent.

### Provenance and peer review

Not commissioned, externally peer reviewed.

### Data availability statement

All data relevant to the study are included in the article. Data are available on reasonable request.

## Notes

### Competing Interest Statement

The authors have declared no competing interest.

### Funding Statement

This work was supported by unconditional donations from Medac, Galapagos and Freunde und Foerderer der Berliner Charite e.V.
This work was supported by grants from COVIM: NaFoUniMedCovid19 (FKZ: 01KX2021) (to L.E.S. and F.Kl.), the Federal Institute for Drugs and Medical Devices (V-2021.3 / 1503_68403 / 2021-2022) (to F.Ku. and L.E.S.).

